# Pain in clients attending a South African voluntary counselling and testing centre was frequent and extensive but did not depend on HIV status

**DOI:** 10.1101/19001784

**Authors:** Antonia L Wadley, Erica Lazarus, Glenda E Gray, Duncan Mitchell, Peter R Kamerman

**Author notes:** Corresponding author: Peter Kamerman, School of Physiology, Faculty of Health Sciences, University of the Witwatersrand, 7 York Rd, Parktown, Johannesburg 2193. Sources of funding: South African National Research Foundation Rated Researchers Programme (GEG).

## Abstract

**Background:** The frequency of pain is reported to be high in people living with HIV (PLWH), but valid comparisons between PLWH and HIV-negative cohorts are rare. We investigated whether HIV infection influenced frequency and characteristics of pain in adults undergoing voluntary testing for HIV.

**Methods:** Participants were recruited from a HIV voluntary counselling and testing (VCT) centre at the Chris Hani Baragwanath Academic Hospital, Soweto, South Africa. Pain was assessed using the Wisconsin Brief Pain Questionnaire. Depressive and anxiety symptomatology was determined using the Hopkins Symptom checklist-25. We then stratified by HIV status.

**Results:** Data from 535 black South Africans were analysed: HIV-infected n=70, HIV uninfected n=465. Overall, frequency of pain was high with 59% (95%CI: 55; 63, n: 316/535) of participants reporting pain, with no difference related to HIV status: HIV-infected 50% (95% CI: 37; 61, n: 35/70), HIV-uninfected 60% (95%CI: 56; 65, n: 281/465). Pain intensity and number of pain sites were similar between the groups as were symptoms of anxiety and depression: mean HSCL-25 1.72 (95% CI 1.57; 1.87) HIV-infected participants and 1.68 (95% CI: 1.63; 1.73) HIV-uninfected participants. Univariate analysis showed female sex and greater depressive and anxiety symptomatology associated with having pain. In a conservative multivariable model, only depressive and anxiety symptomatology was retained in the model.

**Conclusion:** The high frequency of pain found in both HIV infected and uninfected individuals presenting at a VCT centre was more likely to be associated with depression and anxiety, than with the presence or absence of HIV.

## 1. Introduction

The reported prevalence of pain in cohorts of ambulatory people living with HIV (PLWH) ranges from 54-81% compared to the general population prevalence of around 35% (1-5). Possible reasons for the increased prevalence of pain in PLWH are that, in addition to pains that affect the general population, for example, lower back pain and arthritic pain, PLWH have the additional burden of pain directly related to the infection itself, secondary infective and non-infective diseases, and iatrogenic causes (6). Whilst pain severity in PLWH appears to be worse in those with late-stage HIV/AIDS (7, 8), data supporting an association between pain and immunological suppression or viral load are equivocal (5, 6). Thus, treating HIV is not necessarily sufficient to treat the additional burden of pain.

Psychosocial factors such as female sex-at-birth, lower levels of education, depression, sleep disruption, and lower levels of social support (for review see (5)) that are associated with pain in the general population are also associated with pain in PLWH. However, there is little information comparing PLWH to HIV-uninfected people in similar sociocultural and economic conditions, to establish the degree to which physiological, demographic and sociocultural factors contribute to the high pain prevalence in PLWH. One reason for the paucity of information is the difficulty of constructing a control group matching PLWH not just physiologically, socially and demographically, but also psychologically, given the inevitable psychological burden of confirmed HIV infection.

We undertook a prospective cross-sectional study to determine the effect of HIV status on frequency and burden of pain in adults in Soweto, South Africa. Our primary hypothesis was that HIV infection is associated with greater frequency of pain and the secondary hypothesis was that HIV infection increases the overall burden of pain, as assessed by pain intensity, location, and number of pain sites. We also analysed social and demographic factors that might influence the frequency of pain. By assessing pain in clients attending a voluntary counselling and testing centre before they knew their HIV status, we hoped to match the psychological state of the control group to that of PLWH.

## 2. Methods

### 2.1 Setting

Between 19 August 2013 and 20 May 2015 a convenience sample of adult participants of African ancestry was recruited from a HIV voluntary counselling and testing centre (VCT) at the Perinatal HIV Research Unit in Soweto, South Africa. Soweto is located in Gauteng province where HIV infection prevalence at the time was 12.8% (9).

### 2.1 Participants

Clients who were attending the centre for routine HIV testing were eligible if they were 18 years or older, unaware of their current HIV status and able to provide informed consent.

The study was approved by the Human Ethics Research Committee (Medical), University of the Witwatersrand, South Africa (clearance number: M130542). All participants provided written informed consent before undergoing any procedures.

### 2.2 Measures

After providing consent, and before undergoing counselling and HIV testing, participants were interviewed to assess their pain. Trained interviewers used standardized questionnaires originally developed in English and subsequently translated and validated for use in isiZulu and Setswana (10, 11). The consenting process and the interview took place in a private examination room at the testing centre. To account for variations in participants’ ability to self-complete questionnaires, the interviewer asked all the participants questions verbally, and recorded the participant’s answers on printed forms.

#### 2.2.2 Socio-demographic data

The following variables were elicited in the interview: self-declared binary sex (male or female), age (years), education (formal schooling: yes or no, and if ‘yes’, highest school grade and post-school qualification achieved), employment status (full-time employment, part-time employment/piece work, unemployed), and receipt of social grants (currently receiving a grant: yes or no, and if ‘yes’, the type of grant). The outcome of each participant’s HIV antibody tests, and, if they tested positive, CD4 T-cell count, was obtained later from their medical records, with the permission of the participants.

#### 2.2.3 Pain, depression and anxiety assessment

We characterized pain using the Wisconsin Brief Pain Questionnaire (10, 12). The questionnaire assessed the presence and characteristics of pain including distribution and intensity of pain. For the primary outcome - frequency of pain - we identified the participants as having pain, or not, according to how they responded to two questions: i) have you had pain in the last month? ii) Rate your pain at its worst in the past week (assessed on an 11-point numerical pain rating scale anchored at 0=no pain and 10=pain as bad as you can imagine). We classified participants as having pain if they answered “yes”‘ to the first question and they rated their worst pain in the past week as being greater than zero. For pain site distribution, we substituted a tick-box list of body parts potentially affected by pain for the body chart cartoon on the original questionnaire, and we asked participants to identify which affected site was the source of their worst pain.

We assessed depression and anxiety using the Hopkins Symptom Checklist (HSCL)-25, which is a checklist of 15 depression-related and 10 anxiety-related symptoms, with the presence and severity of each symptom in the past week being rated on a 4-point rating scale anchored at 1 = not at all and 4= extremely distressed/bothered (11, 13, 14). Depression and anxiety symptoms were analysed separately and in a combined global score. Scoring involves taking the average score across the items being assessed, so scores range between 1 to 4, with 4 being “extremely distressed/bothered”. We found high levels of correlation between the two dimensions, so we only analysed the global score (Supplement 1).

#### 2.2.4 HIV testing

HIV testing was performed according to the South African National Department of Health guidelines (15). Presence or absence of HIV infection was diagnosed by two point-of-care rapid antibody screening tests employing different test kits. If the results of the rapid tests were discordant, a confirmatory laboratory enzyme-linked immunosorbent assay was performed.

### 2.3 Data analysis

To calculate the sample size required in order to ensure 80% power to detect a difference in the frequency of pain between HIV-infected and HIV-uninfected people, we used a two-sided significance level of 95% with at most a 1:2 recruitment ratio of HIV-infected to HIV-uninfected individuals, based on an estimated 34% prevalence of pain in the HIV-uninfected group (3, 16), and an estimated 56% prevalence of pain in the HIV-infected group (7). These parameters yielded a minimum sample size of 63 HIV-infected individuals and 126 HIV-uninfected individuals.

All data are reported as mean, median, or percentage with a 95% confidence interval. Differences between the HIV-infected and HIV-uninfected groups for frequency of pain, socio-demographic characteristics (e.g., age, schooling, employment), and pain characteristics (e.g., pain intensity, number of pain sites) were assessed using 95% confidence intervals of the difference in mean/percentage. These intervals were interpreted based on whether they included 0, the distance of the lower or upper limit from 0 (when the interval excluded 0), and the width of the interval. All confidence intervals were bootstrap intervals calculated from 999 resamples using the percentile method.

We examined univariate risk factors for having pain using logistic regression with pain/no pain as the dependent variable. We used logistic regression with least absolute shrinkage and selection operator (LASSO) regularization of the coefficients to build the best multivariable model. The penalization parameter (lambda) was selected using 10-fold cross-validation. We built the model under two conditions of lambda, i) with the minimum lambda, and ii) the lambda value within one standard error of the minimum lambda (lambda 1SE). Lambda 1SE penalizes the coefficients more than does the minimum lambda, and so produces a more conservative model (fewer variables and therefore improved interpretability). The advantages of regularized regression methods come at the cost of producing biased estimates, and so we have not reported the regression coefficients for the variable selection component of the analysis. Instead, we present the variables included in each model and the direction of the association. Analysis scripts can be accessed at: https://github.com/kamermanpr/HIV-pain-VCT.git

## 3. Results

Informed consent was completed by 540 individuals. HIV test result data were missing for five participants, thus 535 were included in the final analysis: HIV-infected n=70, HIV-uninfected n=465; recruitment ratio: ∼1:7.6. We achieved the required minimum number of HIV-infected participants and well exceeded the required number of HIV-uninfected participants.

### 3.1 Descriptive data

Descriptive demographic and pain data for the whole cohort and stratified by HIV status are shown in Table 1 (also see Supplements 1 and 2). On average, the HIV-infected group were slightly older than the HIV-uninfected group, and had lower levels of education, though very few participants in either group had fewer than seven years of schooling. There were no differences in pain characteristics between the groups, but the pain burden was high. On average participants had more than one pain site, with the head being the worst affected site, and their worst pain in the past week was, on average, of moderate intensity.

**Table 1.** Characteristics of the cohort

| Variable | Point estimate (95% CI) |  |  |  |
| --- | --- | --- | --- | --- |
|  | Whole cohort<br>N=535 | HIV-infected<br>n=70 | HIV-uninfected<br>n=465 | Difference in<br>mean/frequency HIV+<br>minus HIV- |
| **Demographic information:** |  |  |  |  |
| Age (years, mean)<sup>†</sup> | 34.3 (33.4; 35.3) | 36.8 (34.5; 39.2) | 33.9 (32.9; 35.0) | 2.8 (0.4; 5.5)<sup>‡</sup> |
| CD4 T-cell count (cells/μl, median)<sup>†</sup> | - | 436 (306; 573)- | | - |
| Global Hopkins Symptom Checklist-25 (mean)<sup>†</sup> | 1.68 (1.64; 1.73) | 1.72 (1.57; 1.87) | 1.68 (1.63; 1.73) | 0.04 (-0.12, 0.22) |
| Female (%)<sup>†</sup> | 55 (51; 59) | 64 (52; 75) | 53 (49; 85) | 10 (-2; 22) |
| Schooling (%)<sup>†</sup> |  |  |  |  |
| 0-7 | 5 (3; 6) | 14 (6; 23) | 4 (2; 5) | 11 (3; 20)<sup>‡</sup> |
| 8-12 | 61 (58; 66) | 64 (53; 74) | 61 (57; 66) | 3 (-10; 15) |
| >12 | 34 (30; 38) | 22 (11; 31) | 35 (31; 40) | -14 (-25; -4)<sup>‡</sup> |
| Employment (%)<sup>†*</sup> |  |  |  |  |
| Unemployed | 51 (47; 55) | 57 (46; 69) | 50 (45; 54) | 7 (-5; 20) |
| Employed | 45 (41; 50) | 39 (27; 50) | 47 (42; 51) | -8 (-20; 4) |
| Grant | 4 (2; 5) | 4 (0; 10) | 3 (2; 5) | 1 (-4; 7) |
| **Pain information (only those with pain): n (whole cohort/HIV+/HIV-) = 316/35/281** |  |  |  |  |
| Pain the reason for being tested (%)<sup>†</sup> | 10 (6; 13) | 17 (6; 29) | 9 (5; 12) | 9 (-3; 21) |
| Intensity of worst pain in the past week (0-11 scale) (mean) | 6.8 (6.5; 7.1) | 7.2 (6.3; 7.9) | 6.7 (6.5; 7.0) | 0.4 (0; 1.3) |
| Number of pain sites (median) | 3.5 (3.3; 3.8) | 3.3 (2.7; 4.1) | 3.5 (3.3; 3.8) | -1.0 (-1.5; 1.5) |
| Location of pain (%)<br>(4 most-frequent sites) |  |  |  |  |
| Head | 67 (62; 72) | 63 (46; 77) | 67 (62; 72) | -4 (-21; 12) |
| Low back | 45 (40; 51) | 46 (31; 60) | 45 (40; 51) | 1 (-17; 19) |
| Abdomen | 33 (28; 37) | 29 (14; 43) | 33 (27; 40) | -5 (-21; 11) |
| Chest | 31 (26; 37) | 34 (17; 51) | 31 (26; 36) | 3 (-13; 20) |
| Receiving pharmacotherapy for pain (%) | 46 (41; 52) | 49 (31; 66) | 46 (40; 52) | 3 (-15; 20) |

† Sample size when there are missing data (whole cohort/HIV+/HIV-): Age: n = 532/69/463; CD4 T-cell count<sub>(HIV+ only)</sub>: n = 65; HSCL-25: n = 530/70/460; Female: n = 533/69/464; Schooling: n = 521/70/451; Employment: n = 532/70/462; Pain the reason for being tested: n = 313/35/278
‡ Interval excludes 0; \* Full-time and part-time employed collapsed into single 'employed' group

### 3.2 Primary outcome

The frequency of pain in the full cohort was 59% (n: 316/535; 95%CI: 55 to 63). The frequencies were 50% (n: 35/70; 95%CI: 37 to 61) for HIV-infected and 60% (n: 281/465; 95%CI: 56 to 65) for HIV-uninfected (Figure 1, and Supplement 3). The confidence interval for the difference in the frequency of pain between the two groups was 10% (95%CI: −2.0 to 24). The 95%CI included zero, indicating that the difference in point estimates of the frequency of pain in the two groups was not statistically significant.

**Figure 1.**
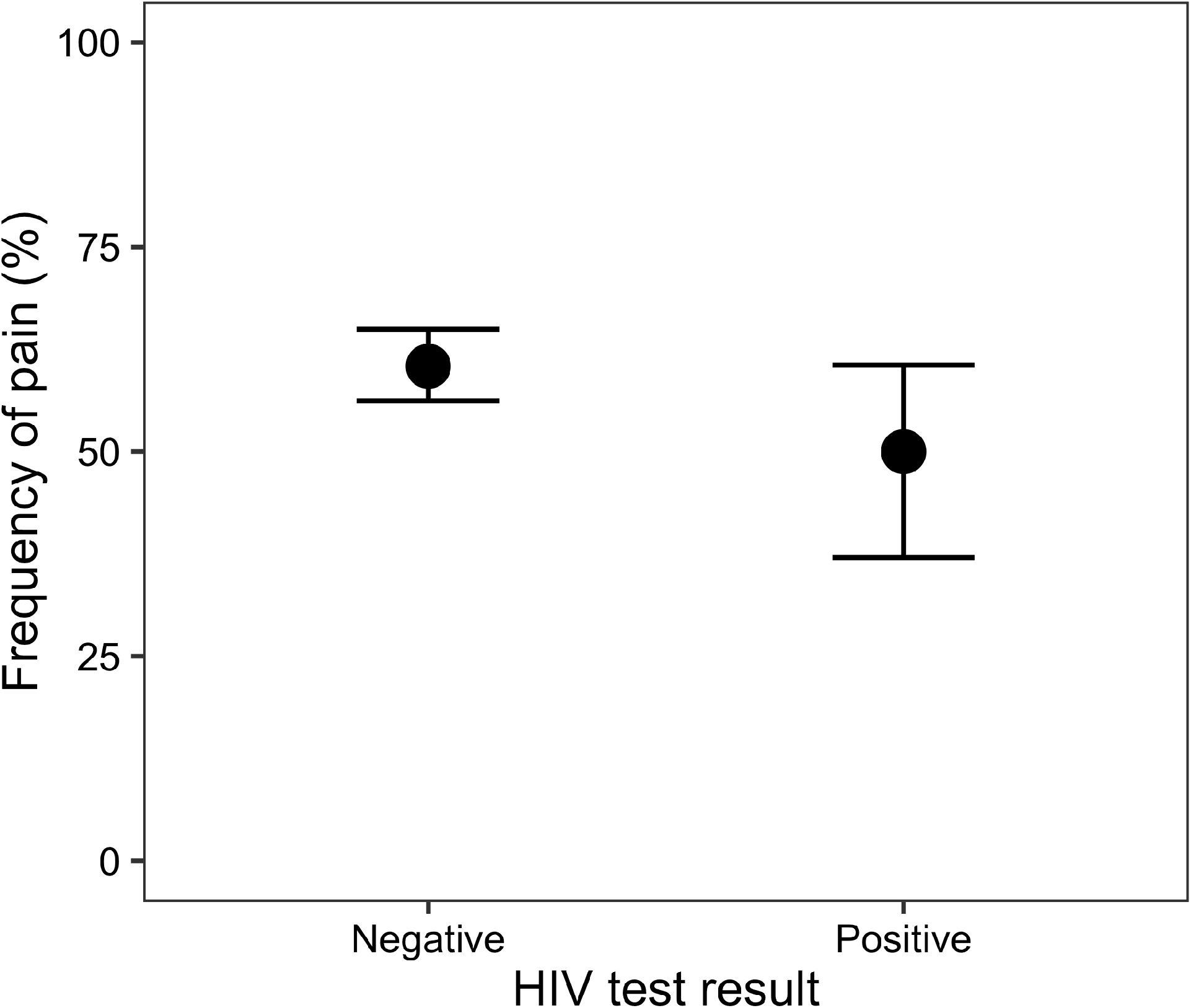
Frequency of pain in participants with (n = 70) and without (n = 465) an HIV infection. The point estimate and 95% confidence interval of the difference in frequency were 10% and −2.0 to 24, respectively.

### 3.3 Risk factors for having pain

Table 2 and Figure 2 (see Supplement 4 for a full description of the models) show the results of logistic regression models for individual predictors for having pain. Based on high levels of correlation between scores for the two dimensions of depression and anxiety we only report the global score (Supplement 1). Of the factors that we investigated, only the total score for the HSCL-25 and being female were significantly associated with having pain. As the HSCL-25 score increased, and hence the severity of anxiety and depression symptomatology increased, likelihood of having pain increased. Similarly, female participants were more likely to have pain than were their male counterparts.

**Table 2.** Results of univariate logistic regression models for the presence of pain.

| *Predictors†* | $\beta$<br>(odds ratio) | $\beta$ 95% CI<br>(odds ratio) | *Likelihood ratio test‡*<br>$\chi^2$ | *df* | *p-value* |
| --- | --- | --- | --- | --- | --- |
| **HIV status** |  |  |  |  |  |
| HIV test results<br>(positive) | -0.423<br>(1.527) | -0.929 to 0.082<br>(0.395 to 1.086) | 2.699 | 1 | 0.100 |
| **Age** |  |  |  |  |  |
| Age | 0.005<br>(1.005) | -0.011 to 0.021<br>(0.989 to 1.021) | 0.390 | 1 | 0.532 |
| **Sex** |  |  |  |  |  |
| Sex<br>(male) | -0.501<br>(0.606) | -0.850 to -0.153<br>(0.427 to 0.858) | 7.988 | 1 | 0.005 |
| **Education** |  |  |  |  |  |
| Level of education<br>(linear) | 0.219<br>(1.245) | -0.400 to 0.828<br>(0.671 to 2.288) | 1.178 | 2 | 0.555 |
| Level of education<br>(quadratic) | 0.028<br>(1.029) | -0.366 to 0.427<br>(0.693 to 1.534) |  |  |  |
| **Employment** |  |  |  |  |  |
| Employment status<br>(employed) | -0.747<br>(0.474) | -1.902 to 0.249<br>(0.149 to 1.282) | 2.245 | 2 | 0.325 |
| Employment status<br>(unemployed) | -0.633<br>(0.531) | -1.786 to 0.360<br>(0.168 to 1.432) |  |  |  |
| **Depression & anxiety** |  |  |  |  |  |
| HSCL-25 total score | 1.367<br>(3.927) | 0.989 to 1.775<br>(2.688 to 5.901) | 59.27 | 1 | 1.4e-14 |

† HIV status levels: Positive/Negative; Sex levels: Male/Female; Education levels (ordinal): grade 0-7/grade 8-12/> grade 12; Employment levels: Grant (pension, child, disability)/Employed (part-time or full time)/Unemployed.
‡ Likelihood ratio test for the overall model.

**Figure 2.**
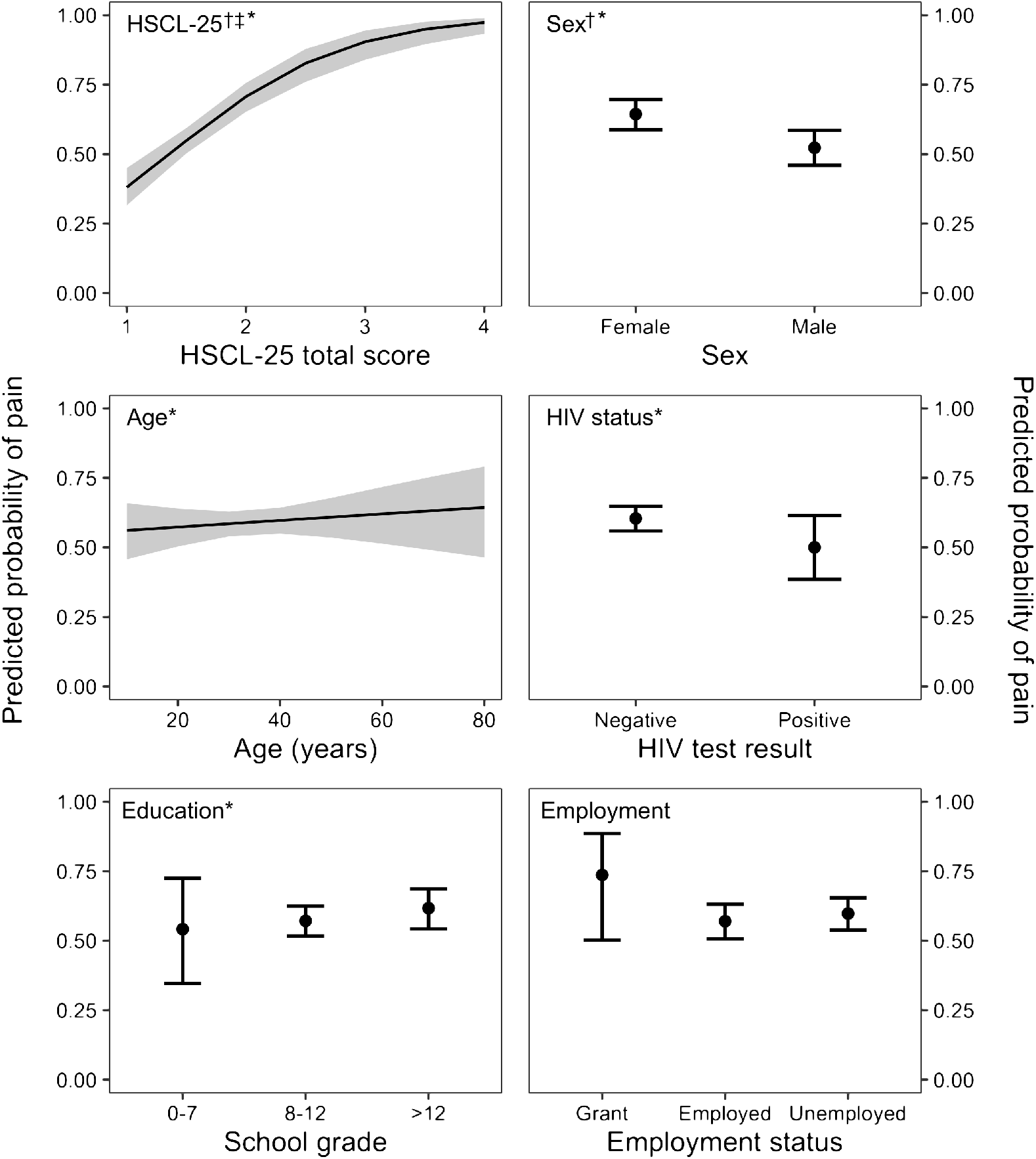
Predicted probability of having pain for six predictor variables (HSCL-25 total score, sex, age, HIV status, education level, and employment status). † Significant on univariable analysis; ‡ Included in a conservative multivariable model; * included in a less conservative multivariable model. See text and Supplement 4 for details of the univariable and multivariable models.

We included all six variables (HIV status, age, sex, education level, employment status, and HSCL-25 score) in a multivariable logistic regression, using LASSO penalised regression for variable selection. Table 3 (Supplement 4) reports the outcomes of the two models that we generated. The model that employed the less conservative lambda minimum retained five of the six variables, such that being HIV-uninfected, having greater HSCL-25 total score, being older, being female were associated with increased likelihood of having pain, and greater levels of education were associated with decreased likelihood of having pain. The more conservative model (that used lambda 1SE) retained only HSCL-25 total score.

**Table 3.**
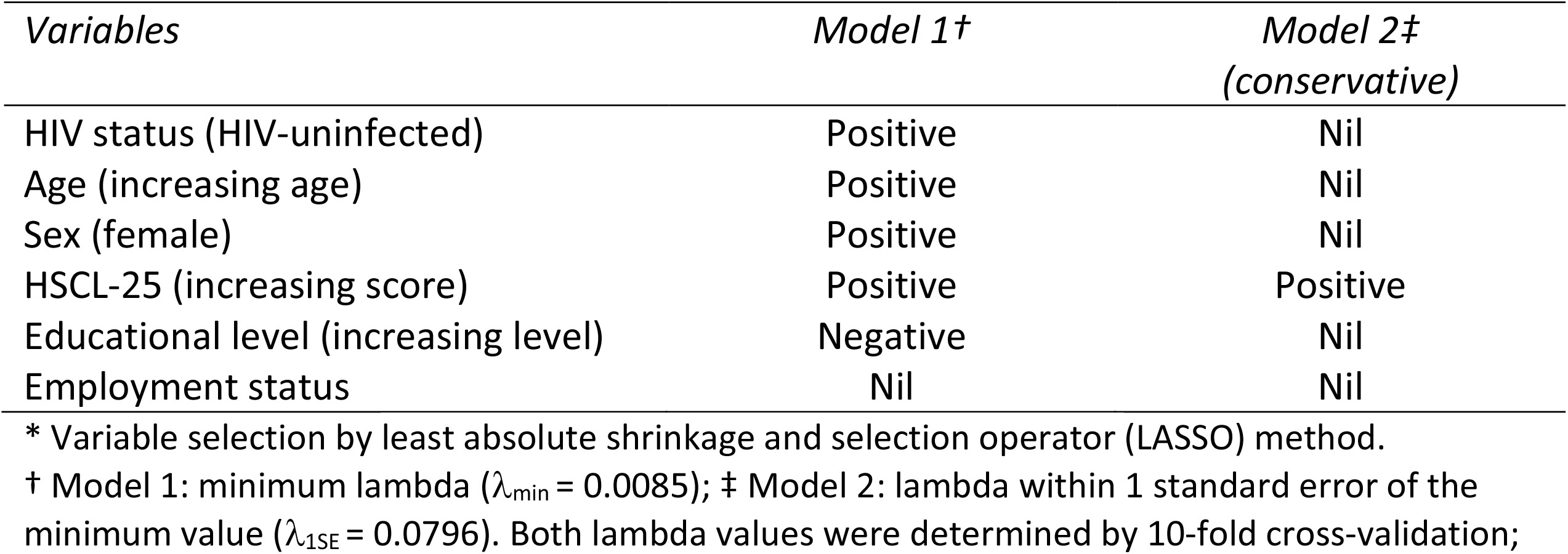
Associations with variables included in multivariable logistic regression models*

## 4. Discussion

The main novelty of our study was that we assessed our hypotheses regarding the effect of HIV infection on the frequency, characteristics and risk factors of pain in a cohort of people whose self-report of pain was not influenced by a prior knowledge of their HIV status. Since participants did not know their HIV status at the time of testing, psychosocial consequences of day-to-day living with HIV, such as dealing with the diagnosis, HIV-related health worries, or enacted or anticipated HIV stigma (17) could not have influenced the development of pain. In addition, since HIV infection had not been diagnosed at the time, HIV-infected participants were not receiving antiretroviral therapy (ART) with their possible algogenic effects. On the other hand, participants who turned out to be HIV-uninfected were sufficiently concerned that they might be infected to have presented voluntarily for testing, so did not have the level of peace of mind that members of a comparison group who knew they were HIV-uninfected might have; such peace of mind may well reduce anxiety and depression, and pain symptoms.

The frequency and extent of pain in the whole cohort was high, irrespective of HIV status, with 59% of the cohort having pain, on average of moderate to severe intensity, and a median of 3.5 pain sites per participant reporting pain. That frequency was in keeping with the frequency of pain in urban ambulatory HIV-infected cohorts in sub-Saharan Africa (7, 18, 19), but well above the frequency in HIV-uninfected comparison cohorts (20, 21), though those comparison groups did not have the attributes of a proper control group. To our knowledge there is only one other published study comparing pain frequency in which there was reasonable matching of attributes in groups of people living with or without HIV (19).

That cohort study, conducted in the UK and Ireland, included a comparison in pain frequency in two groups of people over 50 years of age, one in which the participants were uninfected and the other comprising individuals who were HIV-infected and well-controlled on antiretroviral therapy; the groups were frequency-matched for age, sex, ethnicity, sexual orientation and location (in or out of London). Pain frequency was marginally but significantly higher in the HIV-infected group, at 70%, than in the HIV-uninfected group, at 64%. Participants in the lower-frequency group had the peace of mind of knowing that they were uninfected before their pain was measured. In contrast, our study showed no significant difference in frequency, intensity, location or number of pain sites between those infected and uninfected, in groups matched for age, sex, ethnicity, location and level of employment. Our cohort was younger than were the cohorts of the UK/Ireland study, and our participants with HIV infection were diagnosed at the time of pain testing, implying that they may have had a shorter duration of infection, which could have contributed to the lower pain frequency in our HIV-infected cohort The participants in our HIV-uninfected cohort did not have the benefit of secure knowledge of being free of HIV at the time of pain testing, and nor did our HIV-infected participants know their status at the time of pain measurement, so comparative anxiety levels may have been quite different to those of the cohorts of the UK/Ireland study.

Suggested causes for HIV-related pain include the algogenic effects of the HIV infection itself, pathological processes arising from immune dysregulation as a result of HIV infection, side effects of ART and adjunct therapies, and increased prevalence of depression and sleep disruption (22, 23). Indeed, depression and anxiety, are associated with neuroinflammation, (24) and are common risk factors for pain generally (25). Our data demonstrated more severe depression and anxiety symptomatology to be associated with greater likelihood of pain in both univariate and multivariable analysis, with those participants with the highest scores on the HSCL-25 having a predicted probability of pain close to 100%. Pain and depression are frequently comorbid (26), but this cross-sectional study was not designed to be able to determine the causative relationship between pain and depression, that is whether having pain led to depression, or depression to pain. However, our study did allow us to conclude that the very frequent pain in our cohort of potentially-infected clients presenting at a voluntary counselling and testing centre was associated far more with their depression and anxiety than with the presence or absence of the virus.

In our study, in addition to depression and anxiety symptomatology, female sex was associated with likelihood of having pain, on univariate and less-conservative multivariable analysis. Whilst female sex is a recurring risk factor for pain in general populations (27), whether it is a risk factor in populations of PLWH is equivocal (22, 28, 29), possibly because it has been understudied. Other recurring risk factors for pain, such as increased age, and lower level of education (2, 30), were associated with pain in our cohort on less-conservative multivariable analysis (Table 3), but not on univariate analysis (Figure 2), and the association was not retained statistically on conservative multivariable analysis (Table 3). We conclude that, in our cohort, compared with depression and anxiety, increased age and lower level of education were not key risk factors for pain. The level of education was very similar within the group (most individuals had high school level of education: grades 8 to 12) and the homogeneity may have precluded us from identifying an association. The lack of a strong association with age is surprising, especially considering that ages ranged from less than 20 to more than 80 years. Anomalously, on less-conservative multivariable analysis, being HIV-infected appeared to be protective against likelihood of having pain (Table 3), but this relationship did not appear in univariate analysis (Figure 2) and was not upheld by conservative multivariable analysis (Table 3). Lower socioeconomic status, often associated with lower education and less employment (31), frequently is associated with greater prevalence and intensity of pain (30). Our participants lived in Soweto, a residential area encompassing much low-cost and informal housing. Whilst there may have been a lack of heterogeneity in education, there was a range of individuals employed and unemployed, but state of employment was not associated with pain in any of our statistical analyses. It is possible that even those employed may have been in sufficient poverty for unemployment to have not featured as a risk factor for pain.

We investigated not just the likelihood of having pain but the sites at which pain occurred. Although the head and lower back are commonly the most prevalent pain sites in large pain cohorts, whether HIV-infected or not, the rate of headache in our study was two to three times higher than reported previously for ambulatory PLWH (1, 7, 32). We are unsure of the reason for this discrepancy. A third of participants reported pain in the abdomen and/or chest. This rate of abdominal and chest pain is similar to that in an urban cohort of South Africans living with HIV (7), but those sites did not rank in the top 11 most prevalent pain sites in a survey of 4000 Europeans with chronic pain (1). Many clinicians working in sub-Saharan Africa believe that patients there show somatization of multiple ailments to the chest, and in the Cape Flats, a community similar to Soweto, there was documented somatization of depression as chest pain (33). Indeed, a study of 3000 people from The Netherlands involved in a longitudinal study monitoring depression and anxiety found that 48% of participants had abdominal pain and 26% had chest pain, which was strongly associated with depression and anxiety (34). Furthermore, somatisation may be greater in countries, like South Africa, where ongoing relationships between patient and physician, and thus rapport, are uncommon (35).

There were further limitations to the study. Our study was completed at a single centre in an urban area limiting the generalizability of the results to other populations for example, rural populations. Since we did not record the date of any previous HIV test, it is possible that some individuals who tested HIV-uninfected may have been in the early pre-seroconversion stages of infection. However, since annual HIV incidence in South Africa was estimated as only 1.06% (9), it is unlikely that this failure to detect infection would have had a significant effect on over-reporting of pain in the HIV-uninfected group.

In conclusion, we found that in a high HIV prevalence setting in South Africa, the overall frequency of pain in clients attending a voluntary counselling and testing centre, and assessed for pain before they knew their HIV status, was very high. Pain, however, did not depend on HIV status, or on education level and employment status. However, symptoms of depression and anxiety, which also were very prevalent, and being female sex were associated with likelihood of having pain. Our data do not exclude pain actually associated with the virus, or arising from algogenic effects of treatment, both of which may occur in PLWH for longer than our HIV-infected participants had been. However, our data, the first we believe to include data from properly-matched controls, did reveal a massive contribution to pain frequency in people living with HIV/AIDS from a high burden of pain in the populations from which they are drawn. This pain burden was associated particularly with a high burden of anxiety and depression.

## Data Availability

Study participants did not consent to the public release of their data. However, the data are available on request from Peter Kamerman.

https://github.com/kamermanpr/HIV-pain-VCT.git

